# Modafinil for Wakefulness in the Critical Care Units: A Literature Review and Case Series including COVID-19 Patients at a Tertiary Care Saudi Hospital

**DOI:** 10.1101/2021.02.11.21250832

**Authors:** Marwa Amer, Mohammed Bawazeer, Abid Shahzad Butt, Talal I. Dahhan, Eiad Kseibi, Mouhamad Ghyath Jamil

## Abstract

Cognitive improvement after critical illness is complex. Neuro-stimulants are used to speed up physical and mental processes through the increase in arousal, and wakefulness. In this case series, we reviewed the literature and described the effect of modafinil for wakefulness in a cohort of adult patients admitted to our COVID and non-COVID intensive care unit (ICU) between January 2017 and June 2020. A total of 8 patients included; 3 admitted to COVID-19, 4 surgical, and 1 transplant ICU. Obstructive sleep apnea was noted in 2 (25%), 2 patients (25 %) had an initial neuroimaging that showed hemorrhagic stroke, and 1 (12.5%) showed ischemic stroke with hemorrhagic transformation. Modafinil 100-200 mg daily was started for a median duration of 4 days and the median initiation time in relation to ICU admission was 11 (IQR 9-17) days. Glasgow coma Scale improvement was noted on 5 patients (62.5%). The median duration of mechanical ventilation was 17.5 (IQR 15-31.75) days, and the median ICU stay was 28.5 (IQR 20.25-48) days. All-cause mortality rate was 25 % at 28 days and 62.5 % at 90 days. Modafinil prevented tracheostomy in 1 COVID-19 patient. No significant adverse drug reactions were documented. In our case series, we described our experience for modafinil use for wakefulness in ICU ventilated patients including COVID-19 patients. Based on our observations, the known effects of modafinil, and its safety profile, it holds the potential to facilitate recovery from cognitive impairment. Larger studies are warranted to fully evaluate its role for this indication.

## 1. Introduction

Intensive care unit (ICU) patients are susceptible to neurocognitive and musculoskeletal complications because of various factors, such as the nature of critical illness, medications, over-sedation, and pain [1]. Moreover, sleep disruption in ICUs remains an elusive and pervasive problem. The 2018 pain, agitation/ sedation, delirium, immobility, and sleep disruption (PADIS) guidelines included a brief overview of risk factors contributing to disrupted sleep quality (e.g., architecture, fragmentation, sleep quality score), quantity (e.g., latency, and duration), or circadian rhythm in ICUs [2,3]. Additionally, clinical experience from severe acute respiratory syndrome coronavirus 2 (SARS-CoV-2) suggests its neuro-invasive potential [4-6]. Approximately 36.4% coronavirus disease 2019 (COVID-19) inpatients in a Wuhan hospital exhibited neurological signs and/or symptoms during infection [7]. Manifestations included delirium, confusion, executive dysfunction, anosmia, stroke, headache, Guillain Barré syndrome (GBS), Miller Fisher syndrome, encephalitis, acute necrotizing encephalopathy, myelitis and CNS demyelinating lesions [7,8]. Moreover, critical illness polyneuropathy and ICU-acquired weakness have been observed in 25-46% of ICU patients. Duration of ventilation, corticosteroid administration, multi-organ dysfunction, sepsis, hyperglycemia, and renal replacement therapy have all been correlated with ICU-acquired weakness [9-11]. However, their incidence is unknown in patients with SARS-CoV-2 infection. Case reports of critical-illness polyneuropathy and/or myopathy are rare in patients with SARS-CoV-1 and Middle East respiratory syndrome (MERS-CoV), which can be attributed to the low number of correlation studies between these illnesses and the prolonged ICU stay [12,13].

Early mobilization of these patients has become the standard of care; however, somnolent, fatigued, and depressed patients may not be able or willing to comply and may have a delayed recovery [3]. Neuro-stimulants are used to speed up the physical and mental processes through the increase of dopamine, serotonin, norepinephrine, and acetylcholine. This in turn translates to improved neuro-cognition, enhanced arousal, wakefulness, attention, memory, mental, and motor processing speed [14]. Although, the prescribing trends of neuro-stimulants are yet to be fully elucidated, their use is increasing in traumatic brain injury (TBI) patients especially those with coma, vegetative state (VS), or minimally conscious state (MCS). Neuro-stimulants are sometimes prescribed for off label use, to help in arousal and promote wakefulness in ICUs. The clinical rationale is to facilitate transition from VS, and increase participation in physiotherapy and ambulation, thereby, decreasing ICU-related complications such as deep vein thrombosis (DVT), and ICU-acquired delirium, while facilitating self-expression, accelerating recovery, hastening liberation from the ventilatory support, and decreasing ICU length of stay. Selected neuro-stimulants use in neuro-ICU are summarized at **Table S1** [14-25].

Modafinil is approved by US Food and Drug Administration (FDA) for narcolepsy, obstructive sleep apnea, and shift work sleep disorder. Although the mechanism of action of modafinil remains uncertain, its cognition enhancing properties are attributed to the modulation of glutamate, gamma-aminobutyric acid (GABA) and alpha1B noradrenergic receptors while it has minimal effect on serotonin [26]. Modafinil also offers several advantages over other conventional neuro-stimulants as it does not exhibit the same level of sympathomimetic effects, such as behavioral excitation or rebound hypersomnolence, suggesting that its mechanism of action is distinct. It also has a minimal potential for abuse, and does not appear to disrupt normal sleep architecture. Its favorable safety profile makes it an attractive agent for use in critically ill patients. Side effects include insomnia, anxiety, and cataplexy. At our institution, modafinil is approved for improving wakefulness in adults with excessive daytime sleepiness (EDS) associated with narcolepsy and idiopathic hypersomnia, shift work sleep disorder and as an adjunctive therapy for obstructive sleep apnea (OSA)/hypopnea syndrome. Prescribing privilege is restricted to neurologists, psychiatrists, and sleep medicine consultants. The efficacy of modafinil on wakefulness has been studied in TBI and post-stroke patients [17, 27-32] [**Table 1**]. In ICU setting, a case series by Gajewski and colleagues demonstrated clinical improvement after the start of modafinil therapy in 3 non-intubated ICU patients [31]. A retrospective cohort study by Yoonsun *et al* evaluated whether modafinil improved cognitive function for ICU patients by promoting wakefulness [32]. A total of 60 patients required ventilatory support at a 336-bed community hospital were included. Modafinil resulted in small, nonsignificant increase in Glasgow Coma Scale (GCS) by 0.34 points after controlling for age, baseline severity of illness, and changes in sedation and analgesia over time (95% CI, −0.34 to 0.73 points; p = 0.0743). There was a reduction in exposure to sedatives and analgesics after modafinil was started, resulting in an increasing trend of GCS over time. No major adverse effects were observed. The authors concluded that modafinil did prove beneficial in promoting wakefulness in ICU patients. Its impact on overall patient outcomes in the ICU remains unclear and needs further investigation. In order to gain insight on modafinil use for wakefulness in ICU, we reviewed the literature, described 8 patients, and provided additional clinical data about COVID-19 cases admitted to the ICU. Our aim to describe the clinical characteristics for patients who received modafinil to promote wakefulness and improve cognition, and describe the median change in GCS before and after modafinil therapy. The author also sought to describe the safety, ICU course, and outcomes including duration of mechanical ventilation, ICU length of stay (LOS), hospital LOS, and all-cause of mortality rate.

**Table 1:** Trials on modafinil use in neuro and ICU population

| Study | Patient population | Outcomes | Regimen | Results |
| --- | --- | --- | --- | --- |
| Jha *et al.* Double – blind placebo controlled cross over (2008)/ n= 53 [27] | Moderate – Severe TBI | Fatigue Severity Scale (FSS) and daytime sleepiness (Epworth Sleepiness Scale, ESS) at wk 4 and wk 10 | Modafinil titrated to 400 mg/day or placebo for 10 wk | Clear benefit from modafinil was not seen at wk 4 or wk 10 |
| Kaiser *et al.* Prospective, double blind, randomized, placebo controlled (2010)/ n= 20 [28] | Mild to severe TBI patients in ICU | FSS and ESS at baseline and after 6 wk | Modafinil 100-200 mg or placebo every morning for 6 wk | Modafinil improved excessive daytime sleepiness but not fatigue |
| Poulsen *et al.* Double blinded, placebo controlled (2015)/ n= 41 [29] | Ischemic stroke with modified Rankin Scale score of $\leq 3$ before admission | Fatigue at 90 d (multidimensional fatigue inventory) | Modafinil 400 daily or placebo for 90 d and started 1-14 d post stroke | No significant differences between the 2 groups |
| Bivard A *et al.* Randomized, double blind, placebo- controlled, crossover MIDAS trial (2017)/ n = 36 [30] | Ischemic stroke with multi-dimensional fatigue inventory score of $\geq 60$ | Fatigue (multidimensional fatigue inventory) | Modafinil 200 mg daily or placebo for 6 wk, 1 wk washout, started at least 3 mo post stroke | Modafinil decreased fatigue |
| Gagnon DJ *et al.* systematic review for amantadine and modafinil (2020)/ n= 120 [17] | Post- stroke | Cognitive and functional recovery | Modafinil 170 days post-stroke. Daily doses of 100- 200 mg/day | Modafinil may improve cognitive and functional recovery post-stroke, but higher quality data are needed |
| Gajewski *et al.* Case series (2015) / n=3 [31] | Non-ventilated patients in thoracic surgery ICU | Increase patient wakefulness, and enable a more restful sleep during the night | 200 mg in the first 2 cases and 100 mg in the 3rd case increased later to 200 mg | All 3 pts more engaged in their care and made advances in rehabilitation . No side effect was observed |
| Yoonsun *et al.* retrospective cohort study (2017)/ n= 60 [32] | 60 ICU patients with ventilatory support [medical, surgical, neuro ICU] | Requirements of opioids and sedatives, lowest and average scores of the GCS and SAS during 48 h before and after modafinil | median modafinil dose 170 mg with median duration of 9 d | Modafinil resulted in small, nonsignificant increase in GCS by 0.34 points after controlling for age, baseline severity of illness, and changes in sedation and analgesia over time (95% CI, -0.34 to 0.73 points; p = 0.0743) |
| Current study, observational case series (2020) / n= 8 | 8 ICU intubated patients [3 COVID-19 ICU, 4 surgical ICU, 1 transplant ICU] | Clinical characteristics, median change in GCS 48 h before and 168 h after modafinil | Modafinil 100-200 mg median duration 4 days | GCS improvement was noted on 5 patients (62.5%) |

## 2. Methods

### 2.1 Study design and setting

This was an observational, retrospective, case series performed at a single tertiary care center King Faisal Specialist Hospital and Research Centre (KFSH & RC), Riyadh, the primary referral center in Saudi Arabia. During the COVID-19 pandemic, ICU units were expanded and approximately ~141 COVID ICU beds were added, in addition to approximately ?45 beds in non-COVID ICU units (medical, surgical, transplant/oncology ICUs). The study was approved by the Research Ethics Committee and Clinical Research Committee. The patient’s written consent to publish was taken for each case included in the series. Data were collected and managed using institutional Research Electronic Data Capture (REDCap). The inclusion criteria were 1) ICU patients 18 years and older 2) admitted to COVID and non-COVID units between January 2017 and June 2020 3) ICU stay for at least 48 h, 4) started on modafinil during ICU stay for at least 48 h, and 5) required ventilatory support. Those who did not receive modafinil or received modafinil for indications other than ICU wakefulness were excluded. The CARE (CAse REport) guidelines checklist was followed in describing each case included in this study [33]. The trial registered retrospectively at clinical trial. gov [ClinicalTrials.gov Identifier: NCT04751227]

### 2.2 Variables and statistical approach

Patient demographic data including ICU admission diagnosis, ICU type, comorbidities, and other ICU support [tracheostomy, renal replacement therapy (RRT), steroids, COVID-19 treatment regimen, and vasopressors] were obtained from the electronic medical record (EMR). The status of ventilator use before and after modafinil administration, modafinil initiation time in relation to ICU admission, dose and duration of treatment were also recorded. Severity of illness was evaluated using Sequential Organ Failure Assessment (SOFA) and Acute Physiology and Chronic Health Evaluation (APACHEII) scores. The worst value for baseline laboratory tests was chosen. These data were collected within the first 24 h after admission to the ICU. If that data were not available, then data closest to admission were recorded. Work up for decreased level of consciousness (LOC) was recorded including metabolic abnormalities, neuroimaging [brain computed tomography (CT), and magnetic resonance imaging (MRI)], electroencephalogram (EEG), lumbar puncture, and cerebrospinal fluid (CSF) analysis. The findings of nerve conduction studies (NCS), electromyography (EMG), and other complications during ICU stay were documented. The GCS 48 h before and 168 h (7 days) after modafinil therapy, ICU LOS, hospital LOS, discharge disposition, duration of mechanical ventilation, and mortality rate were recorded. We defined responders for GCS change as a 2 points increase from baseline post modafinil initiation up to 7 days. Duration of mechanical ventilation was recorded as the number of calendar days from intubation to extubation or until ICU discharge, or death whichever occurs first. All-cause mortality rate was reported at 28 and 90 days. The documentation in the EMR was reviewed to identify the patients started on antipsychotics and possible adverse drug reactions (ADRs) related to modafinil including nervousness, agitation, delirium, hypersensitivity, and drug rash. Naranjo scale was used to evaluate the causality of ADRs and modafinil therapy [34].

Continuous variables were presented as mean ± standard deviation (SD) or median (interquartile range [IQR]) as appropriate, and categorical variables were presented as number and frequency (%). The changes in GCS was examined graphically over time. Owing to the retrospective and exploratory nature of this case series, no sample size justification is provided. Instead we wish to report all the patients meeting the inclusion criteria which might be useful to other researchers who may want to combine these results with their own in a meta-analytic approach. Descriptive statistical analysis was conducted using IBM SPSS Statistics, windows version 25.0 (IBM Corp., Armonk, NY).

## 3. Results

### 3.1 Patient characteristics

A total of 8 patients who met the inclusion criteria were described in this case series. The demographics and clinical characteristics are summarized in **Table 2**. The median patient age was 76 (IQR 64.75-80.25) years, 7 (87.5 %) were men, 3 (37.5%) admitted to COVID ICU, and 5 (62.5%) admitted to non-COVID ICU. Regarding the comorbidities, OSA was noted in 2 (25%), and ischemic stroke in 2 (25%). The median SOFA and APACHE II scores were 9.5 (IQR 7.25-11) and 27.5 (IQR 19.5-33), respectively. The median time from ICU admission to modafinil administration was 11 (IQR 9-17) days. Seven patients (87.5%) started on modafinil 100 mg orally every morning while one patient received 200 mg daily for a median duration of 4 days [IQR 3.25-5]. The main indication for modafinil initiation was decreased LOC and altered mental status that potentially preclude successful extubation or liberation from mechanical ventilation. All patients received vasopressors during the ICU stay, and 5 patients (62.5%) started on RRT for pre-existing chronic kidney disease or acute kidney injury. All COVID-19 patients were treated according to our institution protocol for antivirals and immunomodulatory therapies for possibility of cytokine storm. One patient received 5 days of IVIG as part of COVID-19 regimen (patient #1) and one patient received it for possible GBS (patient #8).

**Table 2:**
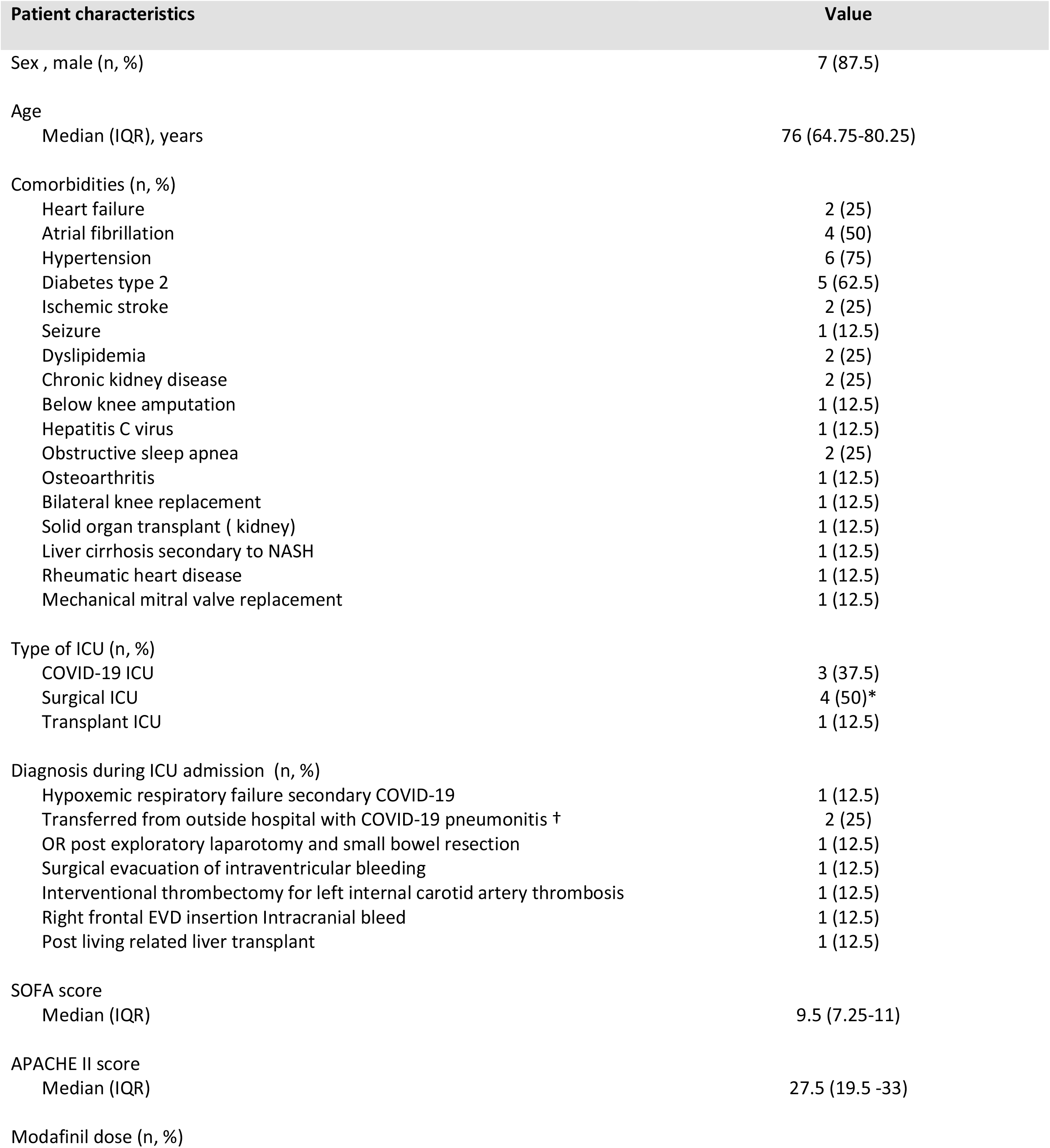
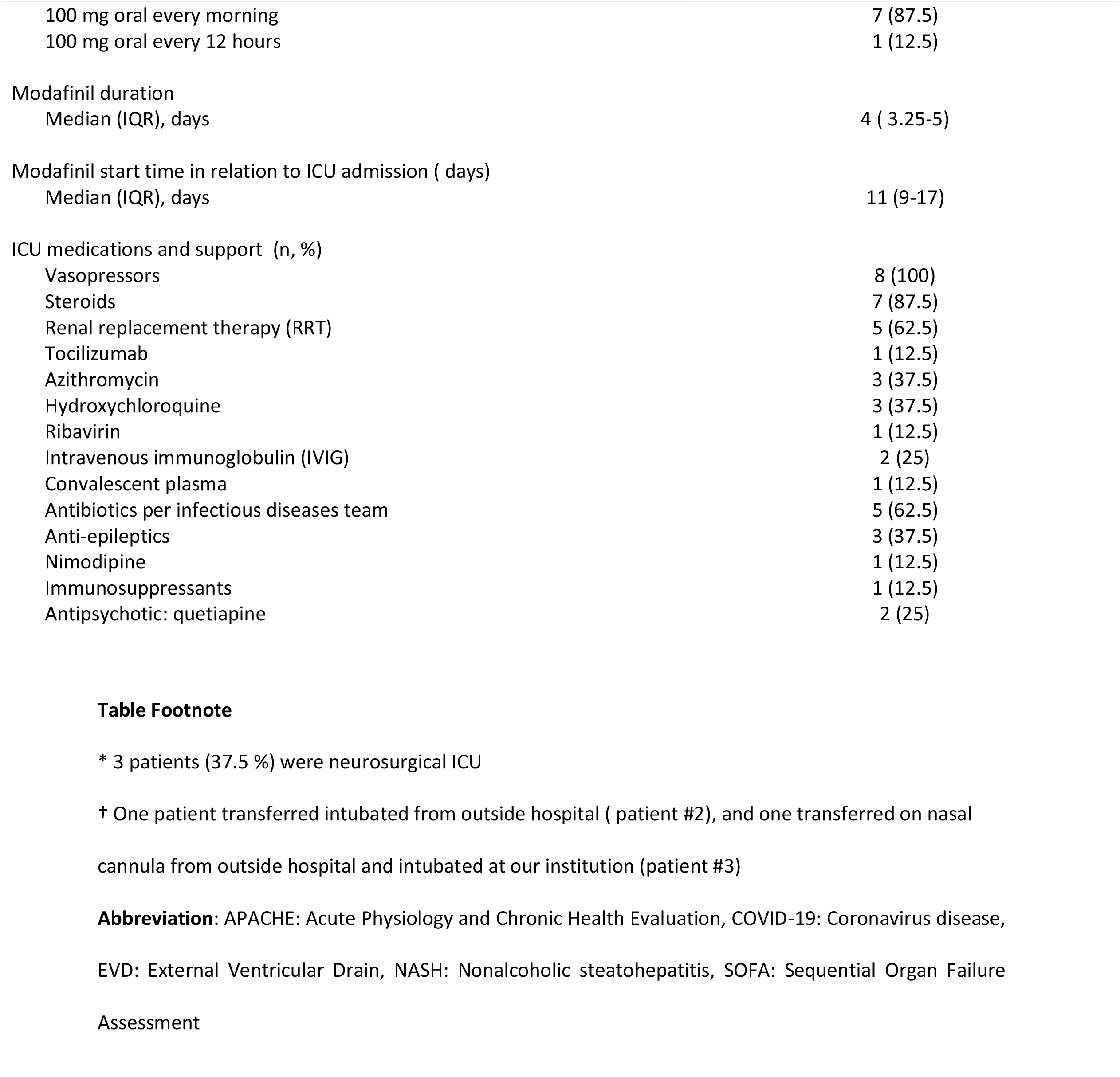
Demographics and baseline characteristics

### 3.2 Follow up and ICU complications

**Table 3** illustrates the workup for decreased LOC during the ICU stay prior to modafinil initiation. Two patients (25%) had initial neuroimaging that showed hemorrhagic stroke, and one (12.5%) had ischemic stroke with hemorrhagic transformation; EEG was performed for 6 (75%) patients and none showed epileptic activity. The cerebrospinal fluid (CSF) analysis was done in 4 patients (50%) and 3 patients showed evidence of nosocomial meningitis that was treated with appropriate antimicrobials [patients # 5-7]. The EMG and NCS were positive in 1 patient (12.5%) for the critical illness myopathy – neuropathy [patient # 8].

Complications arising in the ICU included intra-abdominal infections in 2 (25%) [patients # 4,8], *Clostridium difficile* infection [patients #2,7], endocarditis [patients # 7], cardiac arrest in 2 (25 %) [patients # 6, 8], pulmonary embolism in 1 (12.5%) [patient #2], disseminated intravascular coagulation (DIC) in 1 (12.5%) [patient #2], heparin-induced thrombocytopenia (HIT) in 1 (12.5%) [patient #2], and gastrointestinal bleeding (GIB) and hemorrhagic shock in 3 (37.5%) which required massive transfusion [patients # 2, 4,8]. Antipsychotic drug [quetiapine] was started for 2 patients (25%). No other ADRs were documented.

**Table 3:** workup for decreased level of consciousness (LOC) and complications during ICU stay

| Workup and complications | Value |
| --- | --- |
| Metabolic abnormalities prior to modafinil starts (n, %) |  |
| AKI | 4 (50) |
| ALF | 6 (75) |
| Hypoglycemia | 0 (0) |
| Initial neuroimaging (n, %) |  |
| CT Brain | 6 (75) |
| MRI brain | 2 (25) |
| Time of initial neuroimaging (n, %) |  |
| Day 1 of ICU admission | 3 (37.5) |
| Day 2 of ICU admission | 1 (12.5) |
| Day 4 of ICU admission | 1 (12.5) |
| Day 5 of ICU admission | 1 (12.5) |
| Other * | 3 (37.5) |
| Initial neuroimaging findings | **Patient # 2,3,4:** no evidence of acute intracranial insult<br>**Patient #1:** Stigmata of age related severe microangiopathic ischemic changes with multiple scattered areas of encephalomalacia<br>**Patient #5:** Post-surgical changes from recent evacuation of the right frontal hematoma with extensive IVH and active hydrocephalus<br>**Patient #6:** Status post mechanical thrombectomy with interval development of left basal ganglia and intraventricular hemorrhage. Interval progression of the left MCA territory acute infarction with diffuse hypo attenuation and cytotoxic edema<br>**Patient #7:** evidence of SAH within supratentorial and infratentorial regions with IVH associated with slight interval progression of hydrocephalus<br>**Patient #8:** Small left frontal and right occipital acute/subacute infarcts with associated minimal punctate blood staining in the left frontal lobe. New small bilateral parietal microhemorrhages |
| Repeated neuroimaging (n, %) |  |
| None | 4 (50) |
| CT brain | 3 (37.5) |
| MRI brain | 0 |
| Time of repeated neuroimaging (n, %) |  |
| Day 3 of ICU admission | 1 (12.5) |
| Day 5 of ICU admission | 1 (12.5) |
| Day 7 of ICU admission | 2 (25) |
| Other § | 2 (25) |
| Repeated Neuroimaging findings | **Patient #2:** No CT evidence of acute intracranial abnormality |
|  | **Patient #5:** Status post right frontal craniotomy for evacuation of right frontal IPH and IVH with the placement of two drains. Interval evolution of right frontal IPH and IVH and slight decrease in ventricular dilatation.<br>**Patient #6:** post-surgical changes from decompressive craniectomy and evacuation of basal ganglia hematoma with large left ACA/MCA territory infarction. Interval worsening of the rightward midline shift of 1.1 cm. Interval placement of the left frontal EVD with interval improvement in the size of the right lateral ventricle.<br>**Patient #7 Day 8:** Several probable embolic infarcts in the right centrum ovale and corpus callosum, mild interval improvement of the SAH, no significant interval change of the IVH and ventricular dilatation.<br>**Patient #7 Day 13:** Status post right frontal burr hole for insertion of EVD. No change in ventricular size. Signs of subacute IVH with no evolution of previously noted hemorrhage layering along occipital horns<br>**Patient #7 Day 25:** Interval removal of right EVD, interval decrease of the IVH, with improvement of hydrocephalus, no new hemorrhage or acute abnormality |
| EEG (n, %) | 6 (75) |
| EEG finding of seizure activity (n, %) | 0 (0) |
| Lumbar puncture and CSF analysis (n, %) | 4 (50) |
| EMG and NCS (n, %) | 1 (12.5) |
| EMG and NCS finding: critical illness myopathy – neuropathy (n, %) | 1 (12.5) |
| Complication during ICU stay (n, %) |  |
| Nosocomial meningitis | 3 (37.5) |
| Intra-abdominal infections | 2 (25) |
| Other infections | 5 (62.5) |
| Cardiac arrest | 2 (25) |
| Pulmonary embolism | 1 (12.5) |
| DIC | 1 (12.5) |
| HIT | 1 (12.5) |
| GIB | 3 (37.5) |
| Hemorrhagic shock | 1 (12.5) |

### 3.3 Outcomes

The median change of GCS 48 h before and 168 h after modafinil therapy is displayed in **Fig. 1**. Time 0 the time modafinil was first administered. An improvement was noted in patients # 1, 3, 4, 5, 6. In patient 8, the GCS slightly increased initially after modafinil administration then remains unchanged after that. Patient # 2 and 7, no improvement was noted in GCS after modafinil therapy.

**Figure 1:**
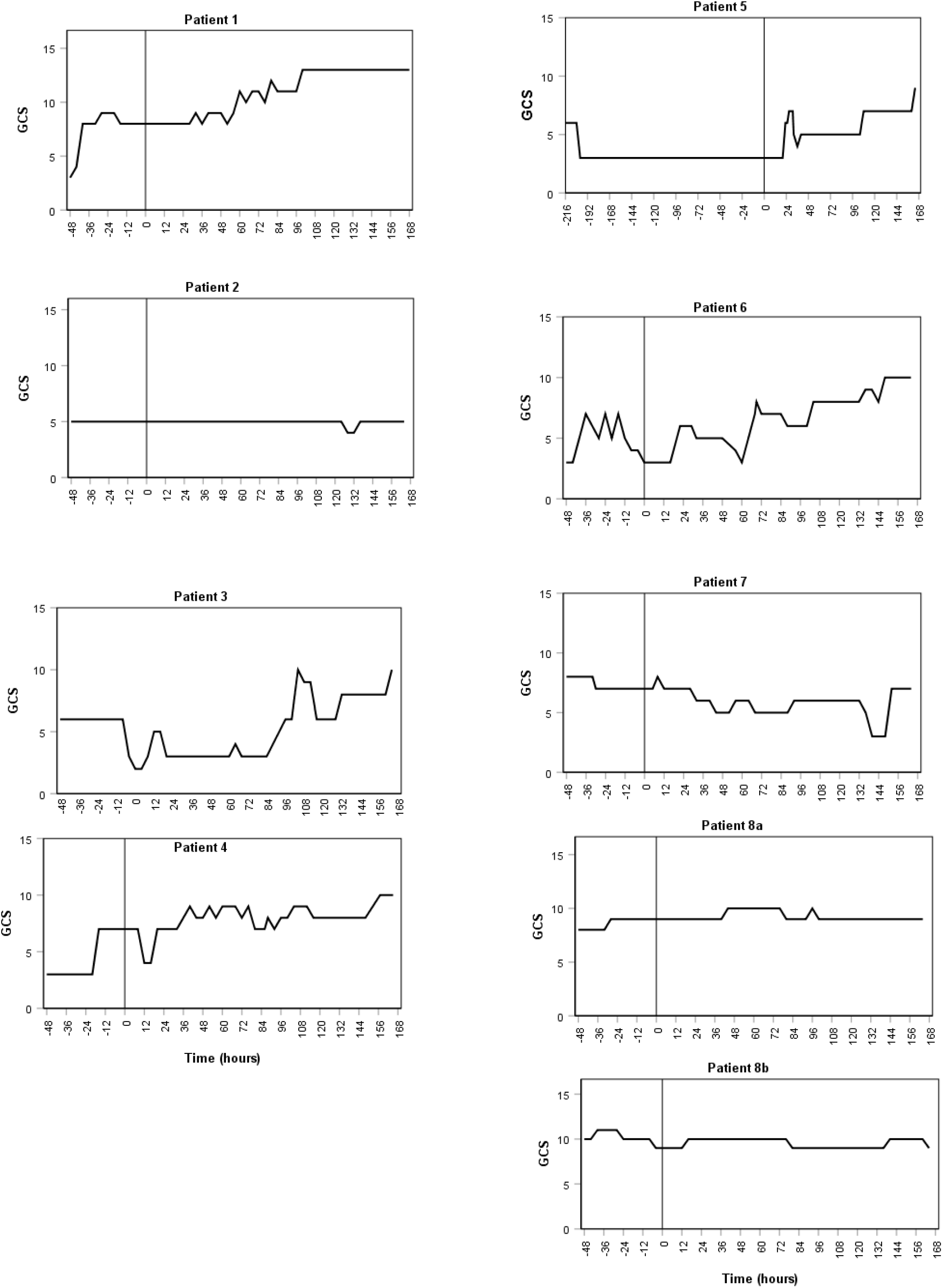
Trend of Glasgow Coma Scale (GCS) over time before and after modafinil

**Table 4** describes the clinical outcomes and patients’ disposition. The median ratio of partial pressure of oxygen and fraction of inspired oxygen (PO2/FiO2) prior to modafinil administration was 223.5 (IQR 175.75-265) which changed slightly after administration, to a median of 195 (IQR 170-390). The median duration of mechanical ventilation was 17.5 [IQR 15-31.75] days; 6 patients (75%) were alive after 28 days and 3 (37.5 %) were alive after 90 days. The median ICU stay was 28.5 (IQR 20.25-48) days, and the median hospital stay was 36.5 (IQR 26.75-115.25) days. Tracheostomy was done in 5 patients (62.5%) and we were able to avoid it in one COVID-19 patient who was subsequently liberated from mechanical ventilation and successfully extubated (patient #1).

**Table 4:** Clinical outcomes and patients disposition

| Clinical outcome | Value |
| --- | --- |
| MV setting prior to start Modafinil |  |
| FIO2 |  |
| Median (IQR) | 0.33 (0.29-0.39) |
| PEEP |  |
| Median (IQR), cmH2O | 8 (5.25-9.5) |
| PaO2/ Fio2 |  |
| Median (IQR) | 223.5 (175.75-265) |
| MV mode (n, %) |  |
| PCV | 2 (25) |
| PS | 2 (25) |
| PRVC | 2 (25) |
| Alternating PCV/PS | 2 (25) |
| A/C | 2 (25) |
| MV setting 48 hours after Modafinil |  |
| FIO2 |  |
| Median (IQR) | 0.30 (0.25-0.40) |
| PEEP |  |
| Median (IQR), cmH2O | 8 (5-9) |
| PaO2/ Fio2 |  |
| Median (IQR) | 195 (170-390) |
| MV mode (n, %) |  |
| PCV | 2 (25) |
| PS | 2 (25) |
| PRVC | 2 (25) |
| Alternating PCV/PS | 1 (12.5) |
| A/C | 0 |
| Duration of MV |  |
| Median (IQR), days | 17.5 (15-31.75) |
| Tracheostomy (n, %) | 5 (62.5) |
| ICU length of stay |  |
| Median (IQR) ,days | 28.5 (20.25-48) |
| Hospital length of stay |  |
| Median (IQR) ,days | 36.5 ( 26.75-115.25) |
| 28-days all-cause mortality (n, %) | 2 (25) |
| 90-days all-cause mortality (n, %) | 5 (62.5) |

## 4. Discussion

In our case series, we described our experience with modafinil for wakefulness in the critical care units including COVID-19 patients at a tertiary care hospital. In majority of the cases, nonpharmacologic measures to encourage a healthy sleep cycle were employed prior to modafinil initiation. Moreover, we addressed multiple potential etiologies for altered mental status prior to modafinil initiation including primary CNS process, metabolic derangements, hypoxemia or hypercarbia due to respiratory failure, renal or hepatic dysfunction, nutritional deficiencies (e.g. thiamine) in patients with poor nutritional status, infection and sepsis induced encephalopathy, medication effects [sedation required for prolonged intubation], EEG to rule out non-convulsive seizure, lumbar puncture if the patient’s neurological findings were not explained by above studies, and NCS to rule out neuromuscular disorders that could preclude successful extubation. Although we cannot state with certainty that modafinil was the primary reason for improvement of cognition function, the temporal relationship between starting modafinil and patients’ improvement makes it a potential contributing factor.

The use of modafinil in a population with neurological findings had been examined before and summarized in table 1 [27-32]. The first trial examined the efficacy of modafinil in treating fatigue and EDS in individuals with TBI [27]. This was a single-center, double-blind, placebo-controlled cross-over trial, where 53 participants were randomly assigned to receive up to 400 mg of modafinil, or placebo. After adjusting for baseline scores and period effects, there were no statistically significant differences between improvements seen with modafinil and placebo on Fatigue Severity Scale (FSS) at wk 4 (–0.5 ± 1.88; p = 0.80) or wk 10 (–1.4 ± 2.75; p = 0.61). For Epworth Sleepiness Scale (ESS) for daytime sleepiness, average changes were significantly greater with modafinil than placebo at wk 4 (–1.2 ± 0.49; p = 0.02) but not at wk 10 (–0.5 ± 0.87; p = 0.56). Modafinil was safe and well tolerated, although insomnia was reported significantly more often with modafinil than with placebo (p = 0.03). Another trial in mild to severe TBI patients in ICU used lower dose of modafinil of 100-200 mg or placebo every morning for 6 wk [28]. This prospective, double-blind, randomized, placebo-controlled pilot study was aimed at studying the effect of daily modafinil on post traumatic EDS and fatigue. The EDS improved significantly in patients with TBI receiving modafinil treatment, compared to the placebo group. Modafinil, however, had no impact on posttraumatic fatigue. Clinically relevant side effects were not observed.

A systematic review for amantadine and modafinil as neuro-stimulants during post-stroke care identified 12,620 publications including 12 modafinil publications (n = 120 patients) [17, 29, 30]. Modafinil was initiated 170 (17, 496) days post-stroke, with initial and final daily doses of 100 (100, 350) mg/day and 200 (100, 350) mg/day, respectively. The most common indication was fatigue for modafinil (n = 5/12; 42%). Only 1 randomized study analyzing the effect of modafinil (8%; n = 21 patients) on acutely hospitalized or ICU patients, witnessed improvement in neurocognitive function. The authors concluded that modafinil may improve cognitive and functional recovery post-stroke, but higher quality data are needed to confirm this conclusion, especially in the acute care setting.

Focusing on ICU setting, Gajewski *et al* published a case series of 3 non-ventilated patients in thoracic surgery ICU without a prior diagnosis of any sleep disorder and receiving a dose of 200 mg modafinil each morning [31]. The primary outcome was to expedite recovery through early rehabilitation. The 1^st^ case was 77-year-old male with a medical history of small non-cancerous lung cells who was readmitted for respiratory distress requiring intubation, then extubated and continued to require intermittent noninvasive ventilation for dyspnea and he was not participating in physical therapy due to hypersomnolence. He was prescribed 200 mg modafinil every morning for 13 d. At day 2, he was weaned on 3-L nasal cannula and made advances in rehabilitation. The 2^nd^ case was a 67-year-old male with a medical history of esophageal cancer. He was admitted to ICU for management of pneumonia and sepsis and received washouts for gastro-pleural fistula. After his initial surgery, he was noted to be extremely fatigued and expressing wishes to withdraw care. Modafinil 200 mg every morning was started and on day 2, the patient was more engaged in his care. No side effects were noted throughout the 8 days course. The 3^rd^ case was a 70-year-old male with a medical history of esophageal cancer was admitted to the ICU for respiratory failure which was improved and eventually extubated. His rehabilitative status was complicated by his overall lack of energy, and his waxing and waning mental status. Modafinil was started at a dose of 100 −200 mg. At day 6, the patient did not need his home lorazepam 1.5 mg at night and he engaged more in his care. The author concluded that modafinil was beneficial in the ICU owing to its effects on improving sleep efficiency, adjusting circadian sleep-cycle disruption, and decreasing EDS which subsequently reduce delirium, improve day-time cognition, improve wakefulness, and facilitate engagement with physical therapy and rehab. Compared to those findings, our observations suggested that modafinil improves cognition and increases GCS in 5 (62.5%) patients in our series, suggesting it might be reasonable for promoting wakefulness in ICU patients especially in mechanically ventilated patients which have not been described in the prior case series [31]. Moreover, we included more diverse cohort from transplant, surgical, and COVID-19 ICUs which had not been reported in the prior literature [31,32]. Mechanically ventilated COVID-19 patients may require higher doses of opioids and sedatives to mitigate ventilator-induced lung injury, and prevent self-extubation. Deep levels of sedation may also require prior to initiation of neuromuscular blocking agents (NMB) in moderate-to-severe acute respiratory distress syndrome (ARDS) [35]. All these factors could potentially delay the recovery of neurocognitive function and increase risks of ICU acquired weakness particularly when corticosteroids are used with aminosteroidal NMB [36]. However, clinicians should also be cognizant that COVID-19 patients are predominately presented with hyperactive delirium, and agitation due to multiple factors including direct virus invasion to CNS, prolonged mechanical ventilation, social isolation, and restricted family visitation [37]. Therefore, comprehensive assessment is warranted to identify those who might benefit from modafinil as wakefulness agent prior to initiation of therapy. Moreover, the most common dose of modafinil that was used in our series 100 mg oral once every morning with a median duration of 4 days and was fairly tolerated. Based on published data, the usual dose of modafinil is 100-400 mg (starting dose 100-200 mg oral daily with a lower dose recommended for patients with cardiovascular diseases and liver cirrhosis) [31, 32]. Therefore, a less ideal response in term of GCS improvement was observed in patients # 2, 7, and 8 which might be due to inadequate modafinil dose. Hence, a higher dose might be reasonable to use in these situations. Regarding the safety and ADRs, we used the Naranjo scale for assessment of causality and a score of 0 to −1 indicated a doubtful causality of ADRs and modafinil therapy in our series [34]. The ICU complications were most likely attributed to underlying comorbidities.

Interestingly, modafinil prevented tracheostomy in one patient with COVID-19 (case 1) who was then successfully extubated. A previous report showed that patients with COVID 19 who required prolonged mechanical ventilation had a high mortality rate. Therefore, tracheostomy might be considered when prolonged mechanical ventilation is anticipated [38]. On the other hand, studies showed that tracheostomy carries significant risk of viral transmission to the healthcare workers [39]. Hence, preventing tracheostomy provided advantages including shortening duration of critical care resource use (ventilators) and ICU length of stay during COVID-19 outbreak, and minimizing aerosolization risks associated with this procedure for healthcare workers. Furthermore, 2 patients (case # 5, and 7) has a neuroimaging that showed hemorrhagic stroke, 1 patient (case #6) showed ischemic stroke with hemorrhagic transformation, a population which had not been assessed for modafinil treatment in the prior literature [17, 27-32]. Moreover, 2 (25 %) of the patients (case #4, and 5) had a history of OSA. We speculated that the observed improvements in cognition and GCS of these patients possibly were related to enhanced nocturnal sleep based on the drug mechanism of action; however, further evaluation is still needed. The 28 d all-cause mortality rate in our series was 2 (25 %) and 90 d all-cause mortality rate was 5 (62.5%) and this is possibly expected as we are a tertiary care hospital. Therefore, it is likely that our patients were sicker, as the APACHEII and SOFA scores suggest that these data were derived from a cohort of critically ill patients. This is comparable to the mortality rate in patients admitted to the ICU with severe sepsis and shock [40-42].

Our study limitations were small sample size, single institution experience, retrospective and observational in nature, and lack of a control group which precludes causal interpretations. Similar to other observational studies, our findings can be biased by confounding from unmeasured factors that influence cognitive function and recovery such as the effect of physiotherapy and rehabilitation. It is also plausible that the increasing trend in GCS over time was largely owing to the result of reduced exposure to other sedatives and analgesics and spontaneous recovery. Moreover, patient-family interaction has a great impact in patient recovery as shown in the recent report [43, 44]. In our series, the intensivist on service had the patient described in case 1 interact with the patient’s family through video calling, which likely uplifted his spirit and played a potential role in his recovery. Additionally, the small sample size prohibited us from performing statistical analysis to assess and adjust for confounders or perform analysis of what factors might be associated with response or lack thereof. This may have led to an overestimation or underestimation of modafinil effect for wakefulness in the critical care unit [45]. As is always the case with observation type study, association does not prove causality, but there are reasonable data showing that modafinil might have potential benefits when administered to certain critically ill patients especially those who appear hypoactive, lethargic, and depressed and particularly when conservative treatment has failed, which provides biological plausibility [17, 27-32]. Its daily cost is insignificant compared with the high cost of an extended ICU stay although it is difficult to conclude from our experience that patients’ length of ICU stay was shortened. Nonetheless, we felt that it is important to report our experience to help rationalize and understand the role of neurostimulants to promote arousal and wakefulness in the ICU which subsequently facilitate self-expression, accelerate recovery, increase participation in physiotherapy and ambulation, hastening liberation from the ventilatory support, and decrease ICU-related complications such as DVT, and ICU-acquired delirium.

Based on our observations of these patients, the known effects of modafinil, its safety profile, and the published experiences of others, we believe that modafinil might has potential benefits and that the consequences of delayed patient recovery and a prolonged ICU stay may outweigh the risks of potential modafinil side effects. Although we cannot state with certainty that modafinil improves cognition function and GCS in our series, it seems that the timing of drug administration in concert with the putative benefits of the medication makes it a plausible conclusion.

There are many unanswered questions that we believe would be a good starting point for a larger randomized controlled trial to substantiate the ideal time of modafinil initiation, identify the population that most likely benefit from therapy, define the optimal drug dose and duration, and identify the role of a combination therapy and its impact on clinical outcomes. Future studies with appropriate sample size, adequate power are needed to better describe its role in the ICU patients post hemorrhagic stroke. Additionally, the impact of modafinil therapy on overall ICU outcomes, such as mortality, duration of mechanical ventilation, and ICU length of stay should be evaluated in future prospective studies. Lastly, further studies are needed to provide more insights and understand modafinil use in critically ill COVID-19 patients. However, in this pandemic setting, evidence of some physiological safety and possible benefit might be considered for hypothesis generation, as shown in our series.

## 5. Conclusion

Cognitive improvement after critical illness is complex. In our case series, we reviewed the literature and described our center’s experience for modafinil use for wakefulness in the ICU ventilated patients including COVID-19 patients. Based on our observations of these patients, the known effects of modafinil, and its safety profile, we believe that modafinil might has potential benefits when administered to certain critically ill patients especially those who appear hypoactive, and lethargic after comprehensive assessment of potential etiologies for altered mental status. Though our observations are intriguing, they result from uncontrolled, retrospective chart review and confounding bias is likely which preclude definitive statements. Nevertheless, and may provide preliminary evidence and hypothesis warrant to be assessed in randomized controlled trials.

## Data Availability

The datasets used and analyzed during the current report are available from the corresponding
author on reasonable request.

## Tables and Figure

**Figure Legend:** The graphs present the median change of GCS 48 h before and 168 h (7 days) after modafinil therapy. The GCS ranges from 3 to 15, with higher scores indicating a higher level of consciousness. Time 0 the time modafinil was first administered. Improvement was noted on patient # 1, 3, 4, 5, 6. No improvement was noted for patient # 2, 7. Patient # 8 received modafinil twice: first trial was 100 mg oral every morning for 5 days [patient 8 a] and the second trial was of 100 mg oral every morning for 6 days [patient 8 b]. In both scenarios, the GCS slightly increased initially after modafinil administration then remains unchanged after that.

**Figure Abbreviation:** GCS: Glasgow Coma Scale

## Supplemental Digital Content

**Table S1** Selected neuro-stimulants use in neuro-ICU

## Acknowledgment

We thank the ICU physicians, ICU nurses, ICU respiratory therapists, ICU physiotherapists, ICU satellite pharmacists, and ICU clinical pharmacists of KFSH&RC for their efforts to save the life of ICU patients and their braveness in fighting against COVID-19 pandemic. We appreciate pharmacy automation team effort and their help to identify the ICU cases for our series [Maher Mominah, and Rania Al-Jaber]. We are also thankful for Hala Khalil, PhD, Department of Biostatistics, Epidemiology and Scientific Computing for providing help in biostatistical part of manuscript.

## Authors contribution

MA: proposed the study, led the manuscript writing and did the data collection. MG: proposed the idea, gave approval for modafinil as a sleep medicine consultant, reviewed and edited the manuscript. MA, MB, AB, MG made substantial contributions to the concept and design of this project and has further participated in the analysis and interpretation of the data. TD, EK: reviewed and edited the final version of the manuscript. All authors participated in the care of the patients, revised the manuscript for important intellectual content and approved the final version to be published. We confirmed that the authorship followed the uniform requirements for manuscripts submitted to biomedical journals.

## Availability of data and material

The datasets used and analyzed during the current report are available from the corresponding author on reasonable request.

## List of Abbreviation

(AKI): acute kidney injury
(ARDS): acute respiratory distress syndrome
(ADRs): adverse drug reactions
(CSF): cerebrospinal fluid
(CT): computed tomography
(COVID-19): coronavirus disease 2019
(CNS): central nervous system
(CRRT): continuous renal replacement therapies
(DVT): deep vein thrombosis
(DM-2): diabetes type 2
(DIC): disseminated intravascular coagulation
(DNR): do-not-resuscitate
(ESRD): end-stage renal disease
(EVD): external ventricular drain
(EEG): electroencephalogram
(EMG and NCS): electromyogram and nerve conduction study
(ED): emergency department
(ESS): epworth sleepiness scale
(EDS): excessive daytime sleepiness
(FSS): fatigue severity scale
(GCS): Glasgow Coma Scale
(GBS): Guillian Barré syndrome
(GABA): gamma-aminobutyric acid
(GIB): gastrointestinal bleeding
(HIT): heparin-induced thrombocytopenia
(HCV): hepatitis C virus
(HFpEF): heart failure with preserved ejection fraction
(HD): hemodialysis
(ICU): intensive care unit
(IVIG): intravenous immunoglobulin
(ICH): intracranial bleeding
(IVH): intraventricular hemorrhage
(KFSH&RC): King Faisal Specialist Hospital and Research Centre
(LOC): level of consciousness
(LP): lumbar puncture
(MERS): Middle East respiratory syndrome
(MI): myocardial infarction
(MCS): minimally conscious state
(MCA): middle cerebral artery
(MRI): magnetic resonance imaging
(NMB): neuromuscular blocking agents
(OR): operating room
(PADIS): Pain, Agitation/ Sedation, Delirium, Immobility, and Sleep Disruption
(RRT): renal replacement therapy
(SARS-CoV-2): severe acute respiratory syndrome coronavirus 2
(SAH): subarachnoid hemorrhage
(TBI): traumatic brain injury
(VS): vegetative state

