## Supplemental table 1 for "Modafinil for Wakefulness in the Critical Care Units: A Literature Review and Case Series including COVID-19 Patients at a Tertiary Care Saudi Hospital"

**Table S1** Selected neuro-stimulants use in neuro-ICU

| **Neuro-stimulant, MOA** | **Study design** | **Study population, N** | **Regimen** | **Results** |
| --- | --- | --- | --- | --- |
| Amantadine- direct and indirect effects on dopamine; weak noncompetitive NMDA receptor antagonist | Meythaler JM *et al* Double blind, randomized, placebo controlled crossover [14] | 35 TBI patients in acute care setting with GCS <10 | Amantadine 100 mg BID started 1-12 wks post injury | Amantadine patients had better functional improvement regardless of the time of amantadine initiation [improvement in the Mini-Mental Status (MMSE) scores of 14.3 points (p=0.0185), Disability Rating Scale (DRS) score of 9.8 points (p=.0022), Glasgow Outcome Scale (GOS) score of 0.8 points (p=.0077)] |
|  | Saniova B *et al* retrospective pilot study [15] | ICU TBI patients with severe head injury (GCS < 8) | Amantadine 200 mg IV, starting on day 3 of hospitalization | Amantadine patients has higher GCS (P < 0.0001) and lower mortality (P<0.0001) than the group treated with standard therapy alone |
|  | Giacino *et al* a multicenter, prospective, double blind randomized controlled trial [16]. | 184 patients with TBI, VS and MCS | Amantadine 100 BID increased to 200 mg BID over 4 wk | During the 4 wk treatment period, recovery was significantly faster in the amantadine group than in the placebo group, as measured by the Disability Rating Scale score (difference in slope, 0.24 points per wk; p = 0.007), indicating a benefit. There were no significant differences in the incidence of serious adverse events |
|  | Gagnon DJ *et al* Systematic review for amantadine and modafinil as neurostimulants [17] | Post‑stroke care | Amantadine was initiated 39 d post-stroke with a dose range 100–200 mg/d | The most common indication was consciousness disorders. A positive response in at least 1 clinical effectiveness measure was reported in 70% of amantadine publications. Most common side effects were visual hallucinations with amantadine (2% of patients) , dizziness (5% of patients) and dry eyes or mouth (5% of patients). |
| Fluoxetine- Selective serotonin reuptake inhibitor (SSRI) | Chollet F *et al* Fluoxetine for motor recovery after acute ischemic stroke (FLAME) study: a multi-center, double-blind, placebo-controlled trial [18]. | 118 ischemic stroke patients with hemiplegia or hemiparesis | Fluoxetine 20 mg PO daily or placebo was given for 3 mo starting 5-10 d after stroke and all received physiotherapy | Improvement in Fugl-Meyer Scale FMMS (d 0–d 90) at d 90 which was significantly greater in fluoxetine group than in the placebo [19·9–28·7]; p=0·003) |
|  | Effects of fluoxetine on functional outcomes after acute stroke (FOCUS): A pragmatic, double-blind, randomized, controlled trial [19]. | 103 hospitals in the UK and included 3127 patients after acute stroke | Fluoxetine 20 mg daily for 6 mo Vs. placebo | Distribution of modified Rankin Scale scores (mRS) at 6 mo did not differ between the fluoxetine and placebo groups. Functional independence (mRS score of 0-2) occurred in 36% of fluoxetine recipients and 38% of placebo recipients, which was a non-significant difference |
|  | Assessment oF FluoxetINe In sTroke recoverY (AFFINITY) trial was a randomized, parallel-group, double-blind, placebo-controlled trial [20] | 43 hospital with acute stroke units. N= 1280 | Fluoxetine 20 mg daily for 6 mo Vs. placebo | The distribution of mRS categories was similar in the fluoxetine and placebo groups (adjusted common odds ratio 0·94, 95% CI 0·76–1·15; p=0·53). Compared with patients in the placebo group, patients in the fluoxetine group had more falls (20 [3%] vs. 7 [1%];p=0·018), bone fractures (19 [3%] vs 6 [1%]; p=0·014), and epileptic seizures (10 [2%] vs 2 [<1%]; p=0·038) at 6 mo. fluoxetine did not improve functional outcome and increased the risk of falls, bone fractures, and epileptic seizure |
| Methylphenidate- Blocks the reuptake of norepinephrine and dopamine | Moein *et al* Randomized double blind [21] | 80 patients with severe TBI (GCS = 5–8) and moderate TBI (GCS = 9–12) | Methylphenidate 0.3 mg/kg/dose ( max 20 mg per dose ) BID vs. placebo | Methylphenidate was associated with reductions in ICU and hospital length of stay by 23% in severe TBI patients (p = 0.06 for ICU and p = 0.029 for hospital stay time). However, in the moderate TBI, there was 26% fall (p = 0.05) only in ICU length of stay |
|  | Johansson et al, Randomized cross over trials on 2014 [22,23]. | Mild to moderate TBI | Methylphenidate (5-60 mg) for 4 wks | Dose- dependent improvement on fatigue and information processing speed (p< 0.05) |
| Amphetamine- increase release of dopamine and norepinephrine from their storage sites and block the reuptake of catecholamines | Cochrane review [24]. | Stroke patients. Ten studies involving 287 patients were included | Variable among the studies | The suggested benefits on motor function and the non-significant trend towards increased risk of death could be related to imbalances in prognostic variables or other bias in the studies. Further research is therefore required |
|  | Double-blind, placebo-controlled trial | 16 patients, during rehabilitation after stroke | randomized to dexamphetamine (10 mg oral) or placebo and physiotherapy | Analysis of variance from baseline to 1 wk follow-up revealed significant improvements in favor of dexamphetamine for subscales ADL (p = 0.023). This small trial suggests that dexamphetamine can augment physiotherapy [25]. |

**Abbreviation**: ADL: Activities of daily living, GCS: Glasgow Coma Score; GABA: gamma-aminobutyric acid; MCS: minimally conscious state; MOA: mechanism of action; NMDA: N-methyl-D-aspartate; PO: *per os;* BID: twice a day; TBI: traumatic brain injury; vs: vegetative state
